# Acute Kidney Injury in Hospitalized Patients with COVID-19

**DOI:** 10.1101/2020.05.04.20090944

**Authors:** Lili Chan, Kumardeep Chaudhary, Aparna Saha, Kinsuk Chauhan, Akhil Vaid, Mukta Baweja, Kirk Campbell, Nicholas Chun, Miriam Chung, Priya Deshpande, Samira S. Farouk, Lewis Kaufman, Tonia Kim, Holly Koncicki, Vijay Lapsia, Staci Leisman, Emily Lu, Kristin Meliambro, Madhav C. Menon, Joshua L. Rein, Shuchita Sharma, Joji Tokita, Jaime Uribarri, Joseph A. Vassalotti, Jonathan Winston, Kusum S. Mathews, Shan Zhao, Ishan Paranjpe, Sulaiman Somani, Felix Richter, Ron Do, Riccardo Miotto, Anuradha Lala, Arash Kia, Prem Timsina, Li Li, Matteo Danieletto, Eddye Golden, Patricia Glowe, Micol Zweig, Manbir Singh, Robert Freeman, Rong Chen, Eric Nestler, Jagat Narula, Allan C. Just, Carol Horowitz, Judith Aberg, Ruth J.F. Loos, Judy Cho, Zahi Fayad, Carlos Cordon-Cardo, Eric Schadt, Matthew A. Levin, David L. Reich, Valentin Fuster, Barbara Murphy, John Cijiang He, Alexander W. Charney, Erwin P. Böttinger, Benjamin S. Glicksberg, Steven G. Coca, Girish N. Nadkarni, on behalf of the Mount Sinai COVID Informatics Center (MSCIC)

## Abstract

**Importance:** Preliminary reports indicate that acute kidney injury (AKI) is common in coronavirus disease (COVID)-19 patients and is associated with worse outcomes. AKI in hospitalized COVID-19 patients in the United States is not well-described.

**Objective:** To provide information about frequency, outcomes and recovery associated with AKI and dialysis in hospitalized COVID-19 patients.

**Design:** Observational, retrospective study.

**Setting:** Admitted to hospital between February 27 and April 15, 2020.

**Participants:** **P**atients aged ≥18 years with laboratory confirmed COVID-19

**Exposures:** AKI (peak serum creatinine increase of 0.3 mg/dL or 50% above baseline).

**Main Outcomes and Measures:** Frequency of AKI and dialysis requirement, AKI recovery, and adjusted odds ratios (aOR) with mortality. We also trained and tested a machine learning model for predicting dialysis requirement with independent validation.

**Results:** A total of 3,235 hospitalized patients were diagnosed with COVID-19. AKI occurred in 1406 (46%) patients overall and 280 (20%) with AKI required renal replacement therapy. The incidence of AKI (admission plus new cases) in patients admitted to the intensive care unit was 68% (553 of 815). In the entire cohort, the proportion with stages 1, 2, and 3 AKI were 35%, 20%, 45%, respectively. In those needing intensive care, the respective proportions were 20%, 17%, 63%, and 34% received acute renal replacement therapy. Independent predictors of severe AKI were chronic kidney disease, systolic blood pressure, and potassium at baseline. In-hospital mortality in patients with AKI was 41% overall and 52% in intensive care. The aOR for mortality associated with AKI was 9.6 (95% CI 7.4-12.3) overall and 20.9 (95% CI 11.7-37.3) in patients receiving intensive care. 56% of patients with AKI who were discharged alive recovered kidney function back to baseline. The area under the curve (AUC) for the machine learned predictive model using baseline features for dialysis requirement was 0.79 in a validation test.

**Conclusions and Relevance:** AKI is common in patients hospitalized with COVID-19, associated with worse mortality, and the majority of patients that survive do not recover kidney function. A machine-learned model using admission features had good performance for dialysis prediction and could be used for resource allocation.

**Key Points:** *Question:* What is incidence and outcomes of acute kidney injury (AKI) in patients hospitalized with COVID-19?

*Findings:* In this observational study of 3,235 hospitalized patients with COVID-19 in New York City, AKI occurred in 46% of patients and 20% of those patients required dialysis. AKI was associated with increased mortality. 44% of patients discharged alive had residual acute kidney disease. A machine learned predictive model using baseline features for dialysis requirement had an AUC Of 0.79.

*Meaning:* AKI was common in patients with COVID-19, associated with increased mortality, and nearly half of patients had acute kidney disease on discharge.

## INTRODUCTION

Preliminary reports indicate that acute kidney injury (AKI) and kidney abnormalities seem to be associated with coronavirus disease (COVID) -19 disease severity and outcomes. A recently published study that utilized autopsy specimens from 26 patients that died of COVID-19 in China,^1^ demonstrated that there is evidence of the invasion of severe acute respiratory distress syndrome coronavirus 2 (SARS-CoV-2) into kidney tissue, along with significant acute tubular injury, endothelial damage, as well as glomerular and vascular changes indicative of underlying diabetic or hypertensive disease.

Small studies from China, Europe and the United States thus far have reported a wide range of the incidence of AKI, ranging between 1 and 42%. The two largest studies published to date show widely disparate rates of AKI. Guan et al. reported an incidence rate of AKI of only 0.5% in 1099 patients from 552 hospitals in China.^2^ The only other report of over 1000 patients with COVID-19 is from Ireland, in which 3908 patients admitted to the ICU in Ireland, Wales and Northern Ireland as of April 24, had a reported incidence of AKI requiring dialysis of 22.2%, with mortality exceeding 75%.^3^

New York City (NYC) has been at the epicenter of the COVID-19 pandemic in the United States and worldwide and as such has had the highest number of hospitalizations. The burden of severe AKI in the setting of COVID-19 has led to widespread shortages in NYC of dialysis nurses, machines, replacement fluids and cartridges for continuous renal replacement therapy (CRRT), and dialysis.^4,5^ The number of AKI cases requiring acute kidney replacement therapy (KRT) has been unprecedented in modern history.

Despite the anecdotal stories regarding the incidence and severity of AKI, the epidemiology of AKI and predictive models for dialysis requirement in hospitalized patients in the United States and particularly NYC, is not well-described. Thus, the objective of this report was to describe the incidence, severity, risk factors and outcomes associated with AKI in the setting of hospitalization of COVID-19 in a major NYC Healthcare System.

## METHODS

### Study Population

The Mount Sinai Health System (MSHS) serves a large, racially and ethnically diverse patient population. In this study, patient data came from the electronic health records (EHR) from five major hospitals that are part of MSHS: Mount Sinai Hospital located in East Harlem, Manhattan; Mount Sinai Morningside located in Morningside Heights, Manhattan; Mount Sinai West located in Midtown and the West Side, Manhattan; Mount Sinai Brooklyn located in Midwood, Brooklyn; and Mount Sinai Queens located in Astoria, Queens. Mount Sinai Beth Israel was not included in this cohort as they used a different EHR.

### Inclusion Criteria

We included patients who were at least 18 years old, had a laboratory-confirmed SARS-CoV-2 infection, and were admitted to any of the aforementioned five MSHS hospitals between February 27 and 12 a.m. on April 15, 2020 (time of data freeze). A confirmed case of COVID-19 was defined by a positive reverse transcriptase polymerase chain reaction (RT-PCR) assay of a specimen collected via nasopharyngeal swab. The Mount Sinai Institutional Review Board approved this research under a broad regulatory protocol allowing for analysis of patient-level data.

### Exclusion Criteria

We excluded patients with known end stage kidney disease (ESKD) prior to admission, patients who were hospitalized for < 48 hours, and patients who were missing laboratory and vital signs during their hospital stay (**Supplemental Figure 1** and **Supplemental Table 1**).

**Figure 1:**
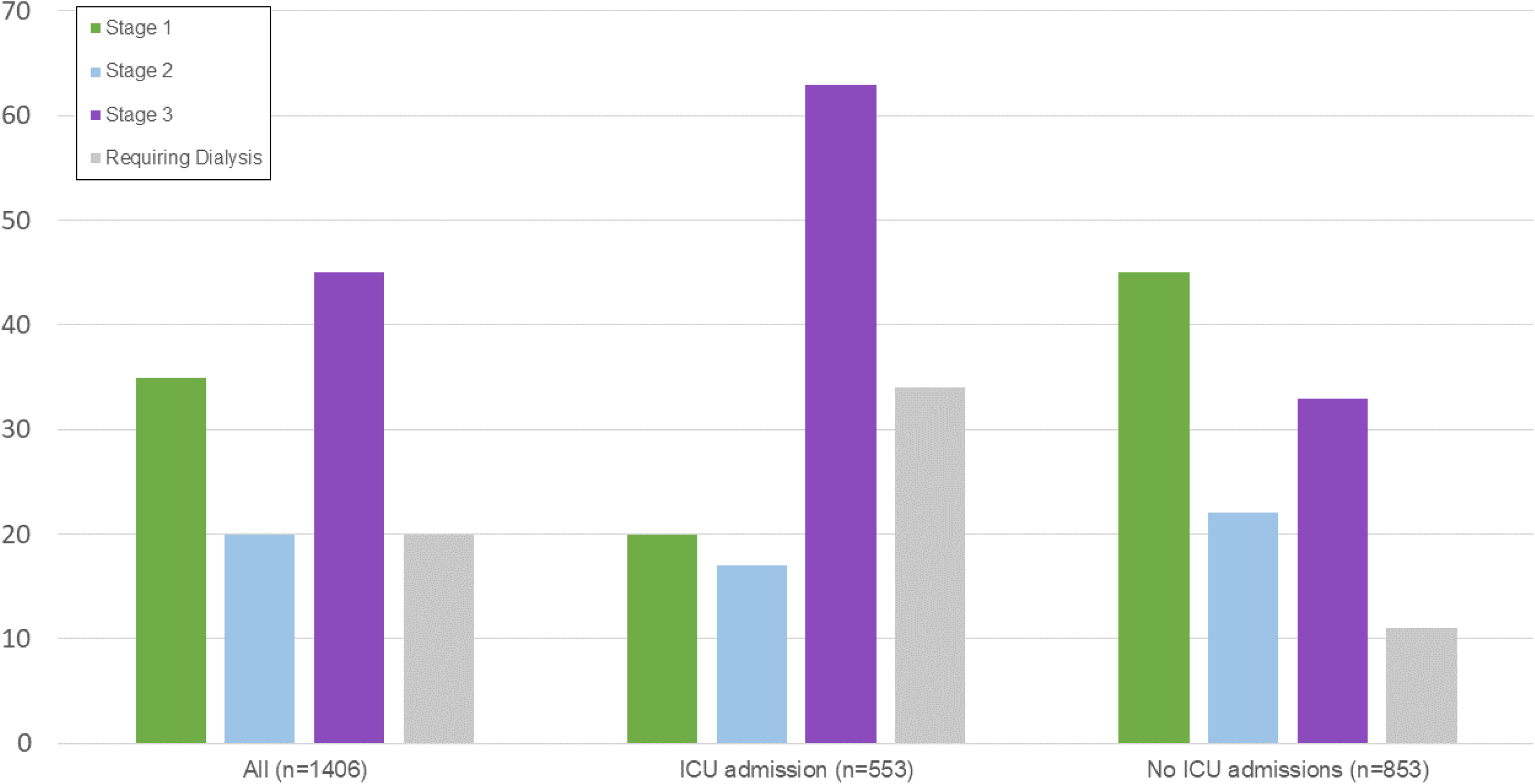
Stages of AKI: Overall and ICU Admissions

### Data Collection

The dataset was obtained from different sources and aggregated by the Mount Sinai COVID Informatics Center (MSCIC); (further description of MSCIC is provided in Supplementary Material). We obtained demographics, diagnosis codes (International Classification of Diseases-9/10-Clinical Modification (ICD-9/10-CM) codes) and procedures, as well as vital signs and laboratory measurements during hospitalization. Demographics included age, sex, and language, as well as race and ethnicity in the EHR. Racial groups included White, Black or African American, Asian, Pacific Islander, Other, and Unknown. Ethnic groups included Non-Hispanic/Latino, Hispanic/Latino, or Unknown. Vital signs and laboratory values during the first 48 hours of admissions that were obtained as part of indicated clinical care were included.

### Definitions of Pre-existing Conditions

We defined a pre-existing conditions as the presence of diagnosis codes associated with specific diseases as per the Elixhauser Comorbidity Software.^6^

### Definitions of Outcomes

AKI, the primary endpoint, was defined by at least an increase in the peak serum creatinine of 0.3 mg/dL or 50% above baseline.^7^ For patients with a previous serum creatinine in the 7 to 365 days prior to admission, the most recent serum creatinine value was considered the baseline creatinine. For patients without a baseline creatinine in the 7 to 365 days prior to admission, the admission creatinine was imputed based upon a Modification of Diet in Renal Disease (MDRD) estimated glomerular filtration fraction (eGFR) of 75 ml/min/1.73m as per the Kidney Disease: Improving Global Outcomes (KDIGO) AKI guidelines.7 We assessed in-hospital mortality defined by survival status at discharge. As a subset of patients was not discharged at the time of this manuscript preparation, subgroup analysis was performed 1) AKI with and without dialysis 2) in patients that were discharged, died, and had at least 14 days, and 3) discharged alive and those who had in-hospital mortality. The need for acute dialysis was ascertained by procedure codes and was cross-referenced with nursing flow sheets. For AKI recovery, we compared the last hospital creatinine with the baseline creatinine and grouped them as recovered, or with acute kidney disease (AKD) Stages 1, 2, or 3 (**Supplemental Table 2**).^8^ We used local regression (loess) for smoothening creatinine values across the interval of two weeks post hospital admission to visualize trajectories by AKD category.

### Statistical Analysis

Baseline characteristics were reported as medians and interquartile ranges or means and standard deviations, for continuous variables. Categorical variables were summarized as counts and percentages. Patients who were still hospitalized at the time of data freeze were regarded as having a censored length of stay (LOS). Logistic regression models adjusted for covariates were used to estimate the adjusted odds ratio for death in patients with AKI vs. without AKI. Covariates were chosen based off of univariate testing and physician input. We used extreme gradient boosting (XGBoost), a boosted decision-tree based machine learning (ML) model, to predict AKI requiring dialysis. Patients from the Mount Sinai (MSH) were randomly split into a training and validation set for the model. To increase model generalizability and help minimize bias, the model’s performance was assessed on a test set composed entirely of patients from the other hospitals (OH) in the MSHS. Features inputted into the model included demographics, laboratory values, and vital signs that occurred in the first 48 hours of admission. For feature importance, we generated SHAP (SHapley Additive explanation) plots where a larger value indicates a larger magnitude of a features effect on a prediction.9 All analyses were performed with R and SAS 9.4.

## RESULTS

A consort diagram of included patients and outcomes is depicted in **Supplementary Figure 1**.

### Demographic and Clinical Characteristics

From February 27 to April 14, 2020, 3235 COVID-19 positive patients who fulfilled our inclusion and exclusion criteria were hospitalized at one of five MSHS New York City hospitals. Baseline creatinine was available prior to admission in 1196 (37%). Patient demographics, pre-existing conditions as well as vital signs and laboratory values stratified by incident in-hospital AKI are displayed in **Table 1**. Patients who developed incident AKI were older and were more likely to have hypertension, congestive heart failure (CHF), diabetes mellitus (DM), and chronic kidney disease (CKD). Patients who developed incident AKI also had higher white blood cell count, lower lymphocyte percentages, higher creatinine values, and higher systolic blood pressure. Other variables were statistically significant but likely not clinically significant given small absolute differences.

**Table 1:** Patient characteristics of all patients and those with and without acute kidney injury

|  | **All (n=3235)** | **AKI (n= 1406)** | **No AKI (n= 1829)** | **p-value** |
| --- | --- | --- | --- | --- |
| **Age Median (IQR)** | 66.5 (55.6-77.8) | 71 (62.5-81.1) | 62 (50.2-73.2) | <0.001 |
| **Sex, n (%)** |  |  |  |  |
| Female | 1367 (42.3) | 579 (41.2) | 788 (43.1) | 0.3 |
| **Race/Ethnicity, n (%)** |  |  |  |  |
| White | 675 (20.9) | 294 (20.9) | 381 (20.8) | <0.001 |
| Black | 702 (21.7) | 355 (25.3) | 347 (18.9) |  |
| Hispanic | 756 (23.4) | 283 (20.1) | 473 (25.9) |  |
| Asian | 82 (2.5) | 30 (2.1) | 52 (2.8) |  |
| Other or unknown | 1020 (31.5) | 444 (31.6) | 576 (31.5) |  |
| **Comorbidities, n (%)** |  |  |  |  |
| Hypertension | 1193 (36.9) | 618 (44) | 575 (31.4) | <0.001 |
| Congestive Heart Failure | 281 (8.7) | 165 (11.7) | 116 (6.3) | <0.001 |
| Diabetes Mellitus | 800 (24.7) | 434 (30.9) | 366 (20) | <0.001 |
| Liver disease | 141 (4.4) | 71 (5.1) | 70 (3.8) | 0.1 |
| Peripheral Vascular Disease | 280 (8.7) | 149 (10.6) | 131 (7.2) | <0.001 |
| CKD | 323 (10) | 243 (17.3) | 80 (4.4) | <0.001 |
| **Lab values, Median (IQR)** |  |  |  |  |
| White blood cell count | 7.5 (5.4-10.6) | 8.5 (5.9-12) | 6.8 (5-9.3) | <0.001 |
| Lymphocyte percentage | 13.1 (8-20.2) | 10.9 (6.6-17.2) | 14.9 (9.2-22.2) | <0.001 |
| Hemoglobin (g/dL) | 12.6 (10.9-14) | 12.5 (10.9-13.9) | 13 (11.8-14.1) | <0.001 |
| Platelets (109/L) | 214 (160-282) | 210 (155-273) | 218 (163-287) | 0.03 |
| Sodium (mEq/L) | 138 (135-141) | 139 (135-142) | 138 (135-140) | <0.001 |
| Potassium (mEq/L) | 4.2 (3.9-4.7) | 4.4 (4-4.9) | 4.1 (3.8-4.5) | <0.001 |
| Chloride (mEq/L) | 103 (100-107) | 104 (100-108) | 103 (100-106) | <0.001 |
| Bicarbonate (mEq/L) | 23.2 (20.8-26) | 22.2 (19.2-25.2) | 24 (22-26.6) | <0.001 |
| BUN blood urea nitrogen (mg/dL) | 18 (12-31) | 31 (19-53) | 13 (10-18) | <0.001 |
| Serum Creatinine (mg/dL) | 0.95 (0.74-1.4) | 1.47 (1.05-2.4) | 0.8 (0.7-0.9) | <0.001 |
| AST (U/L) | 44 (30-71) | 50 (32-83) | 41 (28-62) | <0.001 |
| ALT (U/L) | 31 (19-55) | 30 (18-55) | 32 (20-55) | 0.3 |
| Alkaline phosphatase (U/L) | 75 (54-99) | 77 (60-102) | 73 (57-95) | <0.001 |
| Albumin in (g/dL) | 3 (2.7-3.4) | 2.9 (2.5-3.3) | 3.1 (2.7-3.4) | <0.001 |
| **Vital signs, median (IQR)** |  |  |  |  |
| Temperature (F) | 99.6 (98.6-77.8) | 99.5 (98.6-100.9) | 99.7 (98.7-101.2) | <0.001 |
| Systolic BP (mmHg) | 139 (126-154) | 143 (128-159) | 137 (124-150) | <0.001 |
| Diastolic BP (mmHg) | 80 (73-89) | 81 (73-91) | 80 (73-88) | 0.02 |
| Heart rate (BPM) | 98 (88-110) | 100 (89-114) | 97 (87- 108) | <0.001 |
| Respiratory rate (per min) | 20 (19-25) | 21 (20-28) | 20 (19-23) | <0.001 |
| Oxygen saturation, % | 93 (90-95) | 92 (90-95) | 93 (91-95) | <0.001 |
| **Discharged: n (%)** | 2768 (85.6) | 1124 (80) | 1644 (90) | <0.001 |
| **Hosp. facility** |  |  |  | <0.001 |
| Mount Sinai Hospital<br>located in East Harlem,<br>Manhattan (MSH) | 1317 (40.7) | 505 (35.9) | 812 (44.4) |  |
| Mount Sinai Morningside<br>located in Morningside<br>Heights, Manhattan (STLH) | 600 (18.6) | 274 (19.5) | 326 (17.8) |  |
| Mount Sinai West located in<br>Midtown and the West Side,<br>Manhattan (RVTH) | 408 (12.6) | 143 (10.2) | 265 (14.5) |  |
| Mount Sinai Brooklyn<br>located in Midwood,<br>Brooklyn (BIKH) | 484 (15) | 279 (19.8) | 205 (11.2) |  |
| Mount Sinai Queens<br>located in Astoria, Queens | 426 (13.2) | 205 (14.6) | 221 (14.5) |  |

### Incidence and Severity of AKI

AKI occurred in 1406 patients (43%) and 280 (20%) of those with AKI required dialysis. In the 815 patients admitted to the intensive care, 553 (68%) experienced AKI. Of the 1406 patients with AKI, 40% had AKI at the time of admission. The incidence of AKI in all patients and AKI in the ICU varied by the 5 MSHS locations (**Table 1** and **Supplementary Table 3**). In all admissions, the proportion with stages 1, 2, and 3 AKI were 35%, 20%, 45%, respectively. In those admitted to the ICU, the respective proportions were 20%, 17%, 63%, and 34% required acute renal replacement therapy (**Figure 1**). The median peak serum creatinine was 2.2 (IQR 1.6-3.7) mg/dL in those that did not receive dialysis and was 8.6 (IQR 6.5-11.4) mg/dL in those that did receive dialysis.

In those with baseline creatinine available from the prior 7 to 365 days prior to admission, the overall incidence rate of AKI was 46%, compared to an incidence rate of 42% in those in which the admission serum creatinine was imputed to match an eGFR of 75 ml/min/1.73m2.

### AKI Recovery in Survivors

Of the patients with AKI and who had an outcome (in-hospital mortality or discharge) (n=1,124), 333 (30%) had AKI that had recovered, and 791 (70%) had AKD at the time of discharge (**Figure 2**). The median creatinine at time of discharge or death was 1.9 (IQR 1.1-4.5 mg/dL). In 486 patients with AKI that survived to discharge, 275 (57%) had AKI that had recovered, and 211 (43%) had AKD at the time of discharge; the median serum creatinine at discharge was 1.2 (IQR 0.9-1.6 mg/dL). Recovery from AKI varied by if the patient received dialysis or did not receive dialysis **(Figure 2)**. The characteristics of patients by recovery and each AKD stage is presented in **Supplemental Table 4**. Creatinine trends by recovery group are presented in **Figure 3 A, B and C** for differing subsets of the population.

**Figure 2:**
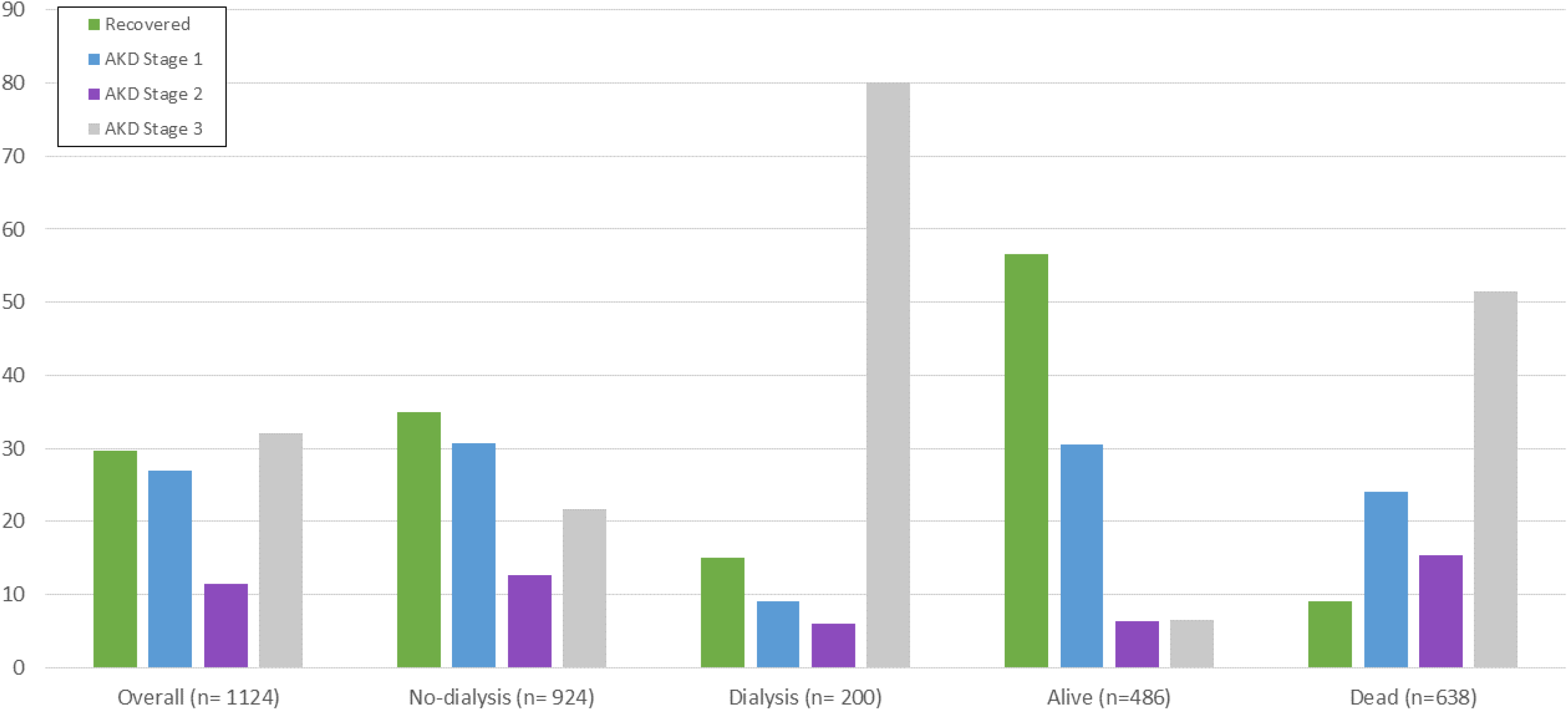
Recovery from AKI in patients who discharged or died

**Figure 3:**
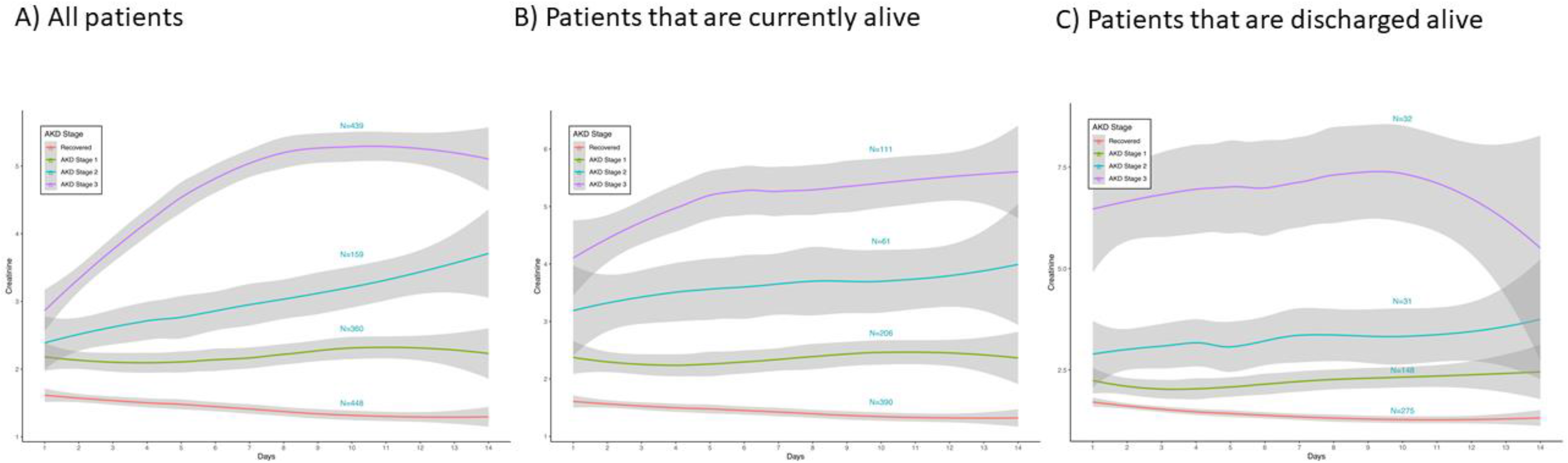
Trajectory of creatinine during the first 14 days of hospitalizations in A) all patients, B) only in patients alive, and C) patients who were discharged alive

### Independent Predictors of Severe AKI

Independent predictors of incident Stage 3 AKI in patients (n=377) who developed AKI while in the hospital (n=848) were CKD (aOR 1.8, 95% CI 1.2-2.7), higher systolic blood pressure (aOR 1.01 per mm Hg, 95% CI 1-1.02), and higher potassium levels (1.3 per 1 meq/L, 95% CI 1.07-1.6) (**Supplemental Figure 2**).

### AKI and Outcomes

Patients with AKI were more likely to have ICU admissions, mechanical ventilation, and use vasopressors administration **(Supplemental Table 5)**. In-hospital mortality in patients that experienced AKI was 45% and was 7% in patients without AKI (P<0.001). In hospital mortality of patients with AKI in the ICU (52%), and non-ICU setting (41%), was markedly higher than those without AKI (ICU 9% and non-ICU 7%). After adjustment for age, gender, race, comorbidities including hypertension, congestive heart failure, diabetes mellitus, liver disease, peripheral vascular disease, CKD, lab values including white blood cell count, leukocyte percentage, hemoglobin, hematocrit, platelets, sodium, potassium, chloride, bicarbonate, BUN, creatinine, AST, ALT, alkaline phosphatase, albumin, vitals including temperature, systolic BP, diastolic BP, heart rate, respiratory rate, oxygen saturation, the adjusted OR for death was 20.9, 95% CI 11.7-37.3 for ICU-AKI vs. no AKI and the adjusted OR for death was 9.6, 95% CI 7.4 – 12.3 for all patients with AKI vs. no AKI (**Supplemental Table 6**).

### AKI Prediction Model

In the training set (n=1,317 patients), the classifier had good performance with an area under the receiver operator curve (AUROC) of 0.79 for predicting AKI requiring dialysis. Performance was similar in the testing set (n=1918) with AUROC = 0.79. **(Supplemental Figure 3)**. The features that had a larger impact on the model included serum creatinine, age, potassium, and heart rate **(Figure 4)**.

**Figure 4:**
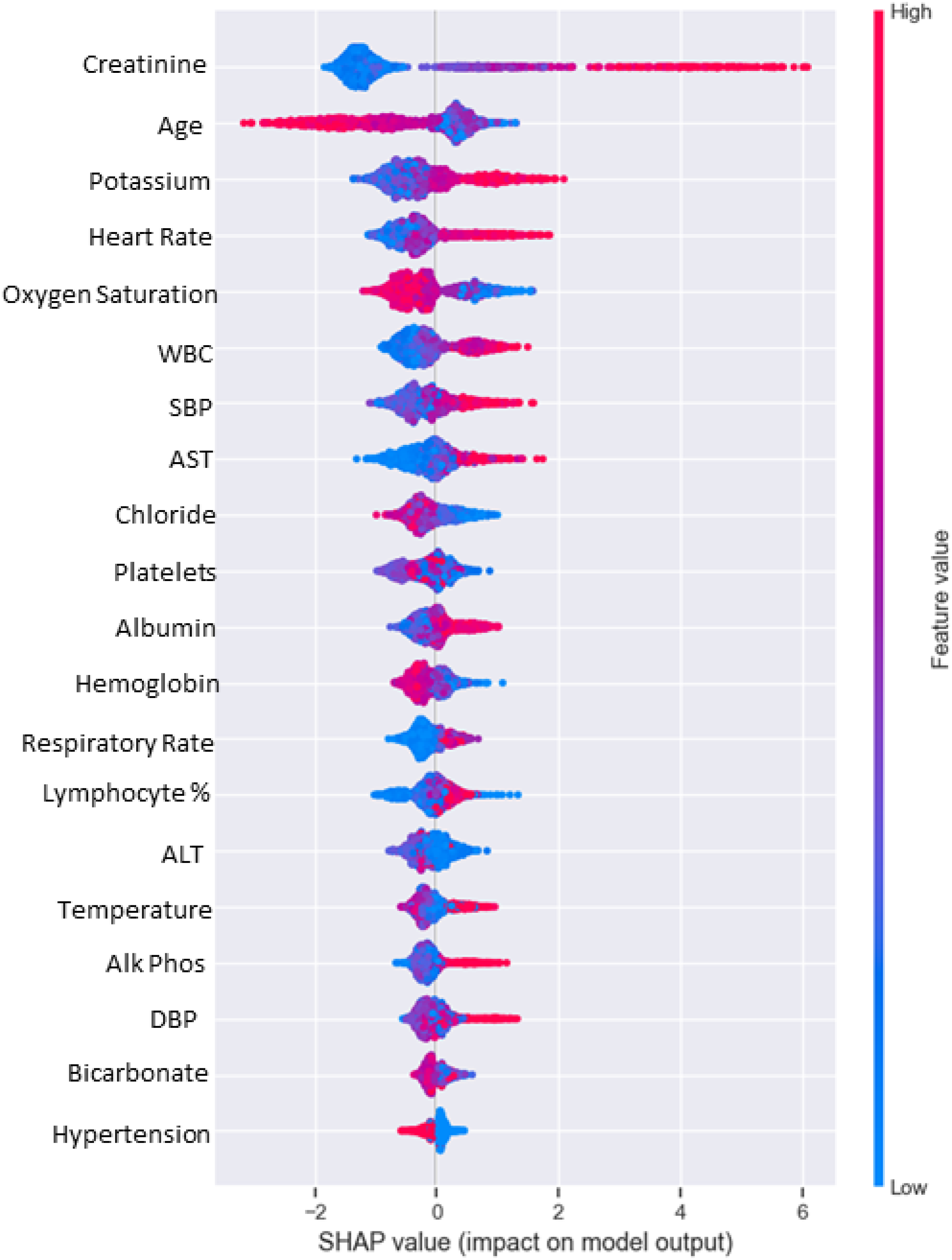
SHAP plots for feature importance

## DISCUSSION

In over 3200 patients with COVID-19 admitted to MSHS, AKI occurred in nearly half of patients and nearly a quarter of those patients required acute dialysis. AKI was independently associated with higher mortality. Many survivors (44%) did not recover kidney function by hospital discharge, and of all patients with AKI, only 30% survived and recovered kidney function. We also trained and tested a machine learning algorithm to predict AKI requiring dialysis with good performance.

While the current COVID-19 pandemic is caused by a coronavirus like the SARS-CoV-2 outbreak in 2005, the reported incidence of AKI associated with COVID-19 disease appears to be substantially higher.^10^ The incidence of AKI in this study is higher than what has been reported in China and Italy and similar to the incidence in another New York City Healthcare system (Northwell).^11^ There are several key differences in the MSHS cohort including higher proportions of patients with comorbidities of hypertension, diabetes, and CKD which are risk factors for developing AKI.^2^ In our analysis, there was no association between hypertension, diabetes with several AKI, but CKD was independently associated with severe AKI. As others have reported, patients who were admitted to the ICU had a higher incidence of AKI likely reflective of more severe disease.

Currently, the exact mechanisms of AKI due to COVID-19 are unclear. A postmortem case series reported by Su et al. found significant acute tubular injury in all their patients.^1^ However, in another case series, in patients with normal creatinine, urinalysis abnormalities (hematuria, proteinuria, and leukocyturia) were common, and unexpected finding in acute tubular necrosis.^12^ Given the high incidence of AKI and lack of full recovery on discharge, identification of potential mechanisms of COVID-19 related AKI would allow for potential interventions to reduce this devastating complication.

This study is the first study in the United States to report the persistence of kidney dysfunction (lack of recovery) in survivors of COVID-associated AKI. Over 40% of patients with AKI in the setting of COVID-19 did not recover kidney function back to baseline. Given the overall severity of AKI as evidenced clinically (high peak creatinine, need for dialysis), as well as the knowledge that many patients with COVID-19 have extensive acute tubular injury on tissue examination, as well as microthrombi, along with a high prevalence of proteinuria, it is not surprising that recovery from AKI was incomplete in most patients.^1^ A study in a Chinese cohort reports similar findings, with less than half of patients fully recovering kidney function.^13^ It remains to be seen what the implications for post-AKI CKD, progression of prior CKD, and ESKD are for COVID-associated AKI.^14-19^ This will require long-term follow-up and further investigation.

Using a small set of features derived early in the hospital admission, we were able to train a ML classifier to predict the need for AKI requiring dialysis with good performance. Several features found to have a big effect on model performance included creatinine, age, and potassium. Additional features such as lymphocyte percentage were found to have more of an impact on the ML classifier than comorbidities such as hypertension. This likely reflects the association of lymphocyte percentage and disease severity as has been previously documented in COVID-19 disease or due to the loss of the protective effects of lymphocytes.^20,21^ Pending additional study of the utility of the ML model, this could be used for resource allocation and planning, to prevent shortages of essential supplies.

Our study should be interpreted in light of the following limitations. Over half of patients did not have a baseline creatinine values. However, we used MDRD imputation as suggested by KDIGO guidelines, and the incidence was nearly identical for those with known baseline serum creatinine vs. those that we imputed because of missing serum creatinine prior to admission.^7^ Recovery was assessed at time of discharge which may not reflect patients’ new baseline kidney function. As the COVID-19 pandemic is still evolving, few long term follow up lab test have been performed. We did not have sufficient data on inflammatory makers (e.g., interleukin 6, ferritin, and fibrinogen) in a large proportion of patients since they were not routinely checked early in the pandemic and a majority of patients did not have these values.^22^ Additionally, urinalysis was also missing in the majority of patients and we did not include in the analysis, although further studies could benefit from characterizing the urinalysis profile of COVID-19 associated AKI. Lastly, baseline medications of interest including angiotensin converting enzyme inhibitors, angiotensin receptor blockers, statins, and other non-steroidal anti-inflammatory drugs were not included.

In conclusion, in a diverse cohort of patients with hospitalized with COVID-19 in NYC, we described a very high incidence of AKI, severe AKI requiring dialysis, and risk of death associated with AKI. We identified several predictors associated severe AKI and found that half of patients did not recover kidney function at time of discharge. Lastly, we developed a ML classifier that had good performance for the prediction of AKI requiring dialysis. The results of this study may be useful to other centers for resource planning during the COVID-19 pandemic, potential additional waves of COVID-19, and for preparing for the increased load of COVID-19 patients with CKD due to severe AKI and lack of recovery.

## Data Availability

N/A

## Acknowledgements

To all the nurses, physicians, and providers who contributed to the care of these patients. To the patients and their family members who were affected by this pandemic.

## Author contributions

Chan, Chaudhary, Coca, and Nadkarni, and members of the MSCIC had full access to all of the data in the study and take responsibility for the integrity of the data and the accuracy of the data analysis.

*Concept and design:* Chan, Coca, Nadkarni

*Acquisition, analysis, or interpretation of data:* Chan, Chaudhary, Coca, Nadkarni

*Drafting of the manuscript:* Chan, Coca, Nadkarni

*Critical revision of the manuscript for important intellectual content:* All authors

*Statistical analysis:* Chaudhary, Chauhan, Saha, Vaid

*Administrative, technical, or material support:* Members of MSCIC

*Supervision:* Coca, Nadkarni

## List of tables and figures

Supplemental Figure 1: Study flow diagram

Supplemental Figure 2: Patient characteristics that were associated with severe AKI (Stage 3)

Supplemental Figure 3: Receiver operate curves for machine learning classifier for outcome of acute kidney injury requiring dialysis of the A) Training and B) Test set.

Supplemental Table 1: ICD codes used for identification of ESKD cohort. ESKD was defined as having both an ESKD diagnosis code and an ESKD procedure code.

Supplemental Table 2: Definitions of acute kidney disease

Supplemental Table 3: Incidence of AKI in patients admitted to the ICU by MSHS hospital, P<0.001)

Supplemental Table 4: Patient characteristics by kidney recovery and acute kidney disease stages.

Supplemental Table 5: Outcomes in patients with AKI and without AKI and the full cohort and in patients who were discharged, had in-hospital mortality, or are admitted with at least 7 days of follow up.

Supplemental Table 6. Adjusted and Unadjusted OR of in-hospital mortality in discharged patients and in ICU admission

